# Adiposity and the risk of dementia: mediating effects from inflammation and lipid levels

**DOI:** 10.1101/2022.03.03.22271843

**Authors:** Ida K. Karlsson, Yiqiang Zhan, Yunzhang Wang, Xia Li, Juulia Jylhävlä, Sara Hägg, Anna K. Dahl Aslan, Margaret Gatz, Nancy L. Pedersen, Chandra A. Reynolds

## Abstract

**Objective:** While midlife adiposity is a risk factor for dementia, adiposity in late-life appears to be associated with lower risk. What drives the associations is poorly understood, especially the inverse association in late-life.

**Methods:** Using results from genome-wide association studies, we identified inflammation and lipid metabolism as biological pathways involved in both adiposity and dementia. To test if these factors mediate the effect of midlife and/or late-life adiposity on dementia, we then used cohort data from the Swedish Twin Registry, with measures of adiposity and potential mediators taken in midlife (age 40-64, n=5999) or late-life (age 65-90, n=7257). Associations between body-mass index (BMI), waist-hip ratio (WHR), C-reactive protein (CRP), lipid levels, and dementia were tested in survival and mediation analyses.

**Results:** One standard deviation (SD) higher WHR in midlife was associated with 25% (95% CI 2-52%) higher dementia risk, with slight attenuation when adjusting for BMI. No evidence of mediation through CRP or lipid levels was present. After age 65, one SD higher BMI, but not WHR, was associated with 8% (95% CI 1-14%) lower dementia risk. The association was partly mediated by higher CRP and suppressed by lower high-density lipoprotein.

**Interpretation:** The negative effects of midlife adiposity on dementia risk were driven directly by factors associated with body fat distribution, and not mediated by inflammation or lipid levels. There was an inverse association between late-life adiposity and dementia risk, especially where the body’s inflammatory response and lipid homeostasis is intact.

## 1. Introduction

Overweight and obesity in midlife is a well-established risk factor for various diseases, including dementia(1). However, when measured in late-life, overweight may instead be associated with lower dementia risk(1), potentially due to reverse causality stemming from unintentional weight loss in the preclinical dementia process(2). While body mass index (BMI) is the most common measure of adiposity, waist-hip ratio (WHR) and other measures of body fat distribution are suggested to be better risk predictors than BMI(3). The negative health effects of higher WHR are hypothesized to stem from central adiposity which indicates increased visceral storage of fat, in and around organs, directly impacting metabolic function. In contrast, gluteal adiposity around the hips indicates subcutaneous fat storage with little metabolic impact(3). Indeed, central adiposity is associated with higher cholesterol levels, insulin resistance, and chronic low-grade inflammation, all of which in turn are associated with dementia(4), and it is possible that these, or other factors, are important players driving the effects of adiposity. In line with this theory, a cross-sectional study of adults aged 20-82 found that WHR is associated with higher burden of white matter hyperintensities in the brain, closely linked to vascular risk factors and associated with higher risk of dementia, and that the effect may be mediated by inflammatory processes(5).

However, in light of the age-specific associations between adiposity and dementia risk, it is plausible that the effects of potential mediating factors also differ between midlife and late-life. Some evidence indicates an inverse association in late-life between what is considered poor cardio-metabolic and dementia risk. Liang and colleagues(6) showed that while good cardiovascular health in midlife was associated with lower dementia risk, the effect was attenuated in late-life. In fact, having good biological cardiovascular health metrics (fasting plasma glucose, total cholesterol, and blood pressure) in late-life was associated with higher dementia risk.

To understand better the age-specific association between adiposity and dementia, we sought to identify and study potential mediating factors. To do so, we first took an empirical approach by using publicly available summary statistics from genome-wide association studies (GWAS) to identify biological pathways shared between adiposity and dementia and which may mediate the association. Secondly, the effects of potential mediators on the associations between BMI, WHR, and dementia were studied in individual level data. In light of the different effects of midlife and late-life adiposity as well as metabolic factors, we examined the risk of dementia in relation to adiposity and biomarker measures taken in midlife and late-life separately. Thus, we aimed to provide a better understanding of what drives not only the association between midlife adiposity and dementia, but also the inverse association between late-life adiposity and dementia.

## 2. Methods

### 2.1 Selection of potential biomarkers through pathway analyses using GWAS summary statistics

To guide the selection of potential biomarker mediators, we used publicly available summary statistics from GWAS to identify biological pathways shared between adiposity and dementia. We used GWAS summary statistics for BMI(7), WHR adjusted for BMI(8) (WHR_adjBMI_), and Alzheimer’s disease(9) (AD). WHR_adjBMI_ was selected instead of WHR, as the goal was to capture any pathways not already included from BMI. The BMI and WHR_adjBMI_ GWAS were both based on around 700,000 individuals, and the AD GWAS on 21,982 individuals with clinically confirmed AD diagnosis and 41,944 controls.

Gene mapping and pathway analysis were carried out in FUMA(10), a web-based tool to assign SNPs to genes, and genes to curated biological pathways. Details on the parameter settings and results are provided in the Supplementary section. For each trait, relevant genes were first identified using the SNP2GENE function. Using the default settings (Table S1), we first identified independent lead SNPs with a GWAS significance of p<5×10^−8^, and candidate SNPs in linkage disequilibrium with the independent lead SNPs with a GWAS significance of p<0.05. The lead and candidate SNPs were then mapped to protein coding genes (excluding the MHC region), based on positional, eQTL, and chromatin interaction mapping(10).

SNPs involved in AD were mapped to 295 genes, BMI to 8663 genes, and WHR_adjBMI_ to 5589 genes (Table S2). Out of these, 175 genes were shared between AD and BMI (Table S3), and 65 genes between AD and WHR_adjBMI_ (Table S4), and selected for pathway analysis.

Pathway analysis was also carried out in FUMA, using the GENE2FUNC function which maps genes to curated biological pathways through enrichment analysis. Using the default setting (Table S5), protein coded genes (n=20,260) were selected as background, and hypergeometric tests used to test if the selected genes (here those in common to AD and BMI or to AD and WHR_adjBMI_) are overrepresented in pre-defined gene sets, after multiple testing corrections. We visually inspected the output plots for GO biological processes in MsigDB c5 to identify potentially mediating biological pathways. The results highlighted biological pathways involved in the immune system and lipid metabolism as common pathways for AD and BMI (Table S6), and lipid metabolism for AD and WHR_adjBMI_ (Table S7).

### 2.2 Study population, individual level data

We used data from four sub-studies of aging in the Swedish Twin Registry (STR)(11). The Swedish Adoption/Twin Study of Aging (SATSA)(12) includes 859 individuals aged 50 and above who participated in up to 10 in-person testing occasions (IPTs) conducted approximately every 3 years between 1986-2014. Aging in Women and Men (GENDER)(13) includes 496 individuals aged 70 and above at baseline who participated in up to 3 IPTs conducted on a 4-year rolling schedule between 1995 and 2005. Origins of Variance in the Oldest Old: Octogenarian Twins (OCTO-Twin)(14) includes 702 individuals who were 80 years or above at baseline and participated in up to 5 IPTs on a 2-year rolling schedule between 1991 and 2002. TwinGene(11) includes 12,630 individuals aged 48-93 who answered a questionnaire and underwent a health checkup between 2004 and 2008. In total, 14,580 individuals participated in one of the sub-studies (some participated in both TwinGene and one of the longitudinal studies).

All participants provided informed consent, and the studies were approved by the Regional Ethical Review Board in Stockholm.

### 2.3 Dementia information

The STR is linked to several nationwide registers, including the National Patient Register (NPR), the Cause of Death Register (CDR), and the Prescribed Drug Register (PDR). From here, dementia information was retrieved(15). Briefly, the NPR includes diagnostic codes (primary and secondary) from all inpatient and specialist outpatient care, including the date of care. The CDR includes main and contributing causes of death. Diagnostic codes used to identify dementia cases were: 304, 305, 306 in ICD-7; 290, 293.0, 293.1 in ICD-8; 290.0, 290.1, 331.0, 290.4, 290.8, 290.9, 294.1, 331.1, 331.2, 331.9 in ICD-9; G30, F01, F02, F03, F05.1, G31.1, G31.8A in ICD-10. Combining dementia information from the NPR and CDR leads to a sensitivity of 63% and a specificity of 99%(16). The PDR includes all dispensed medications since 2005. Medications in the ATC category N06D (anti-dementia drugs Donezepil, Rivastigmine, Galantamine, and Memantine. Tacrine, Ipidacrine, and Ginko folium are not prescribed in Sweden) prescribed through 2016 were used in the current study. In addition, SATSA, OTCTO-Twin, and GENDER entailed a cognitive screening based on the Mini-Mental State Examination (17) and additional cognitive tests as part of the in-person testing. Based on these tests as well as review of medical records, and the research nurse’s evaluation, final dementia diagnoses were determined at multidisciplinary consensus conferences, according to DSM-III-R(18) or DSM-IV(19) criteria.

We designated individuals as having developed dementia if they had either a clinical diagnosis based on information from the in-person evaluation, a diagnostic code for any dementing disorder, or dispensed dementia medication. In the study sample 68% of the dementia diagnoses came from the registers, 13% from diagnoses in SATSA, OCTO-Twin and GENDER, and 19% from both sources. Differential diagnoses were 54% AD, 17% vascular dementia, and 5% mixed pathology of both AD and vascular dementia. As there is a large degree of dementia subtype misclassification in the registers(16), we only examined any dementia in the current study.

### 2.4 Adiposity measures

All IPTs and the TwinGene health checkup included measures of height, weight, waist circumference, and hip circumference. Based on these, BMI was calculated as kg/m^2^, and WHR as waist circumference/hip circumference after carefully cleaning the data(15). For BMI, we excluded values below 15 or above 55, affecting two midlife measures and two late-life measures. For WHR, values above 1.35 were clear outliers and set to missing (>5 standard deviations above the mean, and not consistent with BMI measures), affecting three midlife measures and one late-life measure. Measures were standardized to mean 0 and standard deviation (SD) of 1 prior to analyses, so that the estimates represent the effect of biomarker values one SD above the mean. WHR was standardized separately in men and women.

### 2.5 Biomarker data

Blood samples were collected as part of the IPTs and health checkup. Participants were instructed to fast prior to blood collection, but as fasting was not always feasible information about fasting status was collected. As the immune system and lipid metabolism were highlighted as shared pathways between BMI or WHR_adjBMI_ and AD, the inflammatory marker high-sensitivity C-reactive protein (CRP) and the lipid fractions total cholesterol, high density lipoprotein cholesterol (HDL-c), low-density lipoprotein cholesterol (LDL-c), and triglycerides were selected as potential mediators. CRP, LDL-c, and triglycerides were not available in the OCTO-Twin sample.

Prior to analyses, the distribution of each biomarker was visually inspected. CRP values above 100 were set to missing as it indicates ongoing bacterial infection (n=4 in the midlife sample; and n=4 in the late-life sample). For easier interpretation, all measures were standardized to mean 0 and standard deviation (SD) 1. CRP and triglycerides were strongly skewed, and therefore log-transformed prior to standardization.

### 2.6 Statistical analyses

All analyses were carried out in STATA 16.0(20). Baseline was defined as the first occasion with adiposity and biomarker measures, and individuals were followed from baseline until death or end of register follow-up, with attained age as the underlying time scale. To consider differences between adiposity and potential mediators in midlife and late-life, the sample was divided based on age at baseline, and measures taken in midlife (age 40-64) and late-life (age 65 and above) analyzed separately.

We first examined the effect of adiposity and potential mediators on dementia in Cox proportional hazard regression, modelling risk of dementia in relation to: 1) independent effect of BMI or WHR; 2) joint effects of BMI and WHR; 3) independent effect of each potential mediator; 4) joint effects of BMI or WHR and each potential mediator. All models were adjusted for sex and education (≤7 years or >7 years, corresponding to basic vs more than basic education for these birth cohorts). Models including lipid levels were also adjusted for fasting or not fasting at time of blood sampling. In addition, the strata option was included to allow different baseline hazards across sub-studies, and robust standard errors to account for relatedness of twins. As a sensitivity analysis, steps 1-4 were repeated separately in men and women. To consider a young age at end of follow-up in the midlife sample, a second sensitivity analysis of the midlife sample removed controls younger than 70 at end of follow-up.

Secondly, we performed mediation analyses using the med4way package(21). Mediation analyses test if the exposure affects the mediator, which in turn affects the outcome. For a variable to be formally considered a mediator, there should be an association between the exposure and mediator, and the mediator and outcome(22). To examine mediation and interaction effects, a model for the exposure (BMI or WHR) on the mediator (CRP or lipid levels) is fitted along with a model for the exposure on the outcome (dementia), adjusted for and in interaction with the potential mediator. The total excess risk is then decomposed into four parts as visualized in Figure 1: controlled direct effects of BMI or WHR (explained only by the exposure, and not by the mediator or exposure-mediator interaction), reference interaction (explained only by interaction), mediated interaction (explained by both interaction and mediation), and pure indirect effects (explained only by mediation). A linear regression model was selected for the association between the exposure and mediator, and, since Cox regression can provide biased results in mediation analyses of non-rare outcomes(23), an accelerated failure time (AFT) model with a Weibull distribution was used to estimate the effect of the exposure on the outcome. In contrast to Cox proportional hazard models, which estimate the hazard rate ratio (HRR), AFT models estimate time ratios as an acceleration or deceleration of survival, or disease-free, time. While a HRR above 1 indicates increased risk and below 1 indicates decreased risk of disease, a time ratio above 1 indicates longer disease-free time and below 1 indicates shorter disease-free time. The effect of 1 SD higher BMI or WHR at mean levels of the mediator was tested. If there was evidence of mediation and a significant direct effect of the adiposity measure, the direct effect of adiposity at different levels of the mediator was tested. The models were adjusted for sex, education, sub-study, and, when lipid levels were modelled, fasting status.

**Figure 1:**
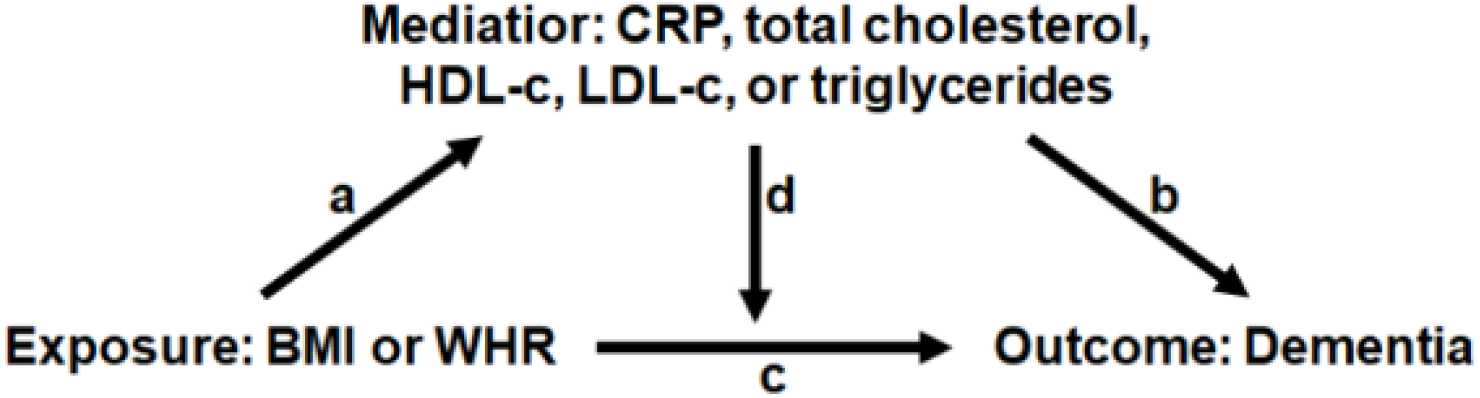
Directed acyclic graph for the relationship between adiposity and dementia, and mediation through CRP or lipid levels. Mediation effects are decomposed into controlled direct effects (through arrow c.), reference interaction (through arrows c. and d.), mediated interaction (through arrows a., c., and d.), and pure indirect effects (through arrows a. and b.).

## 3. Results

### 3.1 Study population

After excluding individuals with no or uncertain dementia information (n=156), younger than 60 at dementia onset (n=6) or last follow-up (n=385), missing covariate information (n=31) or all relevant adiposity and biomarker measures (n=889), or diagnosed with dementia already at baseline (n=220), 12,893 individuals remained for analyses. Out of these, 5999 had midlife measures available and 7257 had late-life measures (363 individuals had measures in both midlife and late-life and were included in both sets of analyses). The midlife sample was followed on average 9.8 years (range 0.6-31.0), during which 110 dementia events occurred, and the late-life sample for 9.0 years (range 0.0-30.6), during which 1000 dementia events occurred. Sample characteristics for the two age groups are presented in Table 1. It should be noted that in the midlife sample age at last follow-up as well as at death was substantially lower among the no dementia compared to the dementia group, mirroring that the midlife sample were followed to a comparatively young age. Indeed, only 7% of the midlife no-dementia sample were deceased at the end of follow-up, compared to 50% of the midlife dementia sample. Removing controls aged younger than 70 from the midlife sample (for sensitivity analyses) left 2,667 individuals for analyses, with more comparative age at end of follow-up (mean 72.7, SD 3.8) and death (mean 78.0, SD 7.0) in the control group.

**Table 1:** Descriptive statistics of the study population.

|  | <u>Age <65 at baseline</u> |  |  |  |  |  | <u>Age ≥ 65 at baseline</u> |  |  |  |  |  |
| --- | --- | --- | --- | --- | --- | --- | --- | --- | --- | --- | --- | --- |
|  | Total sample |  | No dementia |  | Dementia |  | Total sample |  | No dementia |  | Dementia |  |
| Female sex, n (%) | 3353 | (55.89) | 3288 | (55.83) | 65 | (59.09) | 3910 | (53.88) | 3334 | (53.28) | 576 | (57.60) |
| Lower education, n (%) | 1435 | (23.92) | 1394 | (23.67) | 41 | (37.27) | 2985 | (41.13) | 2509 | (40.10) | 476 | (47.60) |
| Age at baseline, mean (SD) | 59.09 | (3.90) | 59.06 | (3.91) | 60.44 | (3.32) | 72.13 | (5.94) | 71.60 | (5.77) | 75.43 | (5.93) |
| Age at last follow-up, mean (SD) | 68.95 | (5.19) | 68.77 | (4.96) | 78.54 | (7.42) | 81.60 | (6.20) | 80.97 | (6.01) | 85.54 | (5.93) |
| Age at death, mean (SD) | 73.50 | (8.36) | 72.52 | (8.01) | 81.39 | (6.81) | 84.53 | (6.97) | 83.85 | (7.14) | 86.81 | (5.83) |
| Dementia onset, mean (SD) | 74.89 | (7.21) | -- | -- | 74.89 | (7.21) | 82.35 | (6.24) | -- | -- | 82.35 | (6.24) |
| BMI, mean (SD) | 25.95 | (3.96) | 25.95 | (3.97) | 26.14 | (3.29) | 25.98 | (3.93) | 26.04 | (3.95) | 25.62 | (3.77) |
| WHR, mean (SD) | 0.88 | (0.08) | 0.88 | (0.08) | 0.92 | (0.08) | 0.89 | (0.08) | 0.89 | (0.08) | 0.89 | (0.08) |
| CRP, mean (SD) | 2.62 | (4.42) | 2.62 | (4.41) | 2.89 | (6.02) | 3.42 | (5.79) | 3.50 | (6.06) | 2.86 | (3.33) |
| Total cholesterol, mean (SD) | 5.87 | (1.11) | 5.86 | (1.10) | 6.54 | (1.31) | 5.89 | (5.89) | 5.87 | (1.21) | 6.02 | (1.23) |
| HDL cholesterol, mean (SD) | 1.43 | (0.42) | 1.42 | (0.42) | 1.52 | (0.43) | 1.42 | (1.42) | 1.42 | (0.42) | 1.43 | (0.42) |
| LDL cholesterol, mean (SD) | 3.81 | (0.96) | 3.81 | (0.96) | 3.91 | (1.15) | 3.74 | (3.74) | 3.74 | (1.01) | 3.74 | (1.02) |
| Triglycerides, mean (SD) | 1.33 | (0.82) | 1.33 | (0.82) | 1.45 | (0.83) | 1.45 | (1.45) | 1.44 | (0.82) | 1.50 | (0.81) |

Descriptive statistics for individuals with measures taken in midlife (age 40-64) and late-life (age 65 and above), for the total sample and by dementia status.
Statistics are presented as number of individuals (%) for categorical variables or mean level (SD) for continuous variables. BMI: body mass index; CRP: C-reactive protein; HDL: high-density lipoprotein; LDL: low-density lipoprotein; N: number; SD: standard deviation; WHR: waist-hip ratio

### 3.2 Midlife measures of adiposity and potential biomarkers in relation to dementia

#### 3.2.1 Independent and joint effects of adiposity, CRP, and lipid levels on the risk of dementia

In the full midlife sample, 1 SD higher WHR was associated with 1.25 times higher risk of dementia (95% CI 1.02-1.52), while 1 SD higher BMI was associated, though not significantly, with 1.13 times higher risk (95% CI 0.96-1.33). Including both adiposity measures in the same model slightly attenuated the estimates to an HRR of 1.20 (95% CI 0.97-1.49) for WHR and 1.10 (95% CI 0.91-1.33) for BMI. None of the biomarkers assessed in midlife were significantly associated with dementia risk, and adjusting for the biomarkers did not substantially affect the BMI or WHR estimates (Table 2a-b).

**Table 2:** Independent and joint effects of adiposity and potential mediators on the risk of dementia.

|  | CRP | Total cholesterol | HDL cholesterol | LDL cholesterol | Triglycerides |
| --- | --- | --- | --- | --- | --- |
| **a) <u>Midlife BMI and potential mediators</u>** |  |  |  |  |  |
| N with both measures | 5,396 | 5,996 | 5,955 | 5,529 | 5,996 |
| **Independent effect models** |  |  |  |  |  |
| BMI → dementia | 1.16 (0.95-1.42) | 1.13 (0.96-1.33) | 1.08 (0.92-1.26) | 1.14 (0.93-1.40) | 1.13 (0.96-1.33) |
| Mediator → dementia | 0.87 (0.63-1.21) | 1.16 (0.98-1.38) | 1.13 (0.91-1.40) | 1.09 (0.78-1.51) | 1.09 (0.89-1.34) |
| **Joint effect models** |  |  |  |  |  |
| BMI → dementia, adjusted for mediator | 1.27 (0.99-1.62) | 1.12 (0.95-1.32) | 1.14 (0.97-1.35) | 1.14 (0.92-1.41) | 1.12 (0.94-1.33) |
| Mediator → dementia, adjusted for BMI | 0.79 (0.55-1.15) | 1.15 (0.97-1.37) | 1.18 (0.95-1.48) | 1.09 (0.79-1.52) | 1.05 (0.85-1.31) |
| **b) <u>Midlife WHR and potential mediators</u>** |  |  |  |  |  |
| N with WHR and biomarker measure | 5,311 | 5,802 | 5,765 | 5,441 | 5,803 |
| **Independent effect models** |  |  |  |  |  |
| WHR → dementia | 1.27 (0.96-1.68) | **1.24 (1.01-1.52)** | 1.18 (0.96-1.45) | 1.28 (0.98-1.66) | **1.25 (1.02-1.52)** |
| Mediator → dementia | 0.92 (0.66-1.28) | 1.21 (0.98-1.48) | 1.07 (0.85-1.36) | 1.13 (0.81-1.58) | 1.11 (0.90-1.36) |
| **Joint effect models** |  |  |  |  |  |
| WHR → dementia, adjusted for mediator | **1.33 (1.00-1.76)** | **1.24 (1.00-1.53)** | 1.22 (0.97-1.52) | 1.28 (0.98-1.67) | 1.23 (0.98-1.53) |
| Mediator → dementia, adjusted for WHR | 0.85 (0.59-1.20) | 1.20 (0.98-1.47) | 1.13 (0.89-1.44) | 1.13 (0.81-1.58) | 1.05 (0.85-1.30) |
| **c) <u>Late-life BMI and potential mediators</u>** |  |  |  |  |  |
| N with BMI and biomarker measure | 6,470 | 7,252 | 6,810 | 6,061 | 6,891 |
| **Independent effect models** |  |  |  |  |  |
| BMI → dementia | **0.93 (0.86-1.00)** | **0.92 (0.86-0.99)** | **0.91 (0.84-0.98)** | **0.90 (0.83-0.98)** | **0.93 (0.86-0.99)** |
| Mediator → dementia | **0.90 (0.84-0.97)** | 0.98 (0.92-1.05) | 0.96 (0.89-1.04) | 1.03 (0.95-1.11) | 1.04 (0.97-1.11) |
| **Joint effect models** |  |  |  |  |  |
| BMI → dementia, adjusted for mediator | 0.95 (0.88-1.02) | **0.92 (0.86-0.98)** | **0.89 (0.82-0.96)** | **0.90 (0.83-0.98)** | **0.91 (0.84-0.98)** |
| Mediator → dementia, adjusted for BMI | **0.92 (0.85-0.99)** | 0.98 (0.92-1.04) | 0.93 (0.86-1.01) | 1.02 (0.95-1.11) | 1.07 (0.99-1.14) |
| **d) <u>Late-life WHR and potential mediators</u>** |  |  |  |  |  |
| N with both measures | 6,416 | 7,121 | 6,687 | 6,005 | 6,760 |
| **Independent effect models** |  |  |  |  |  |
| WHR → dementia | 1.02 (0.95-1.10) | 1.01 (0.94-1.08) | 1.00 (0.93-1.07) | 1.01 (0.94-1.09) | 1.02 (0.95-1.09) |
| Mediator → dementia | **0.90 (0.84-0.97)** | 0.98 (0.91-1.04) | 0.97 (0.89-1.04) | 1.03 (0.95-1.11) | 1.03 (0.97-1.11) |
| **Jont effect models** |  |  |  |  |  |
| WHR → dementia, adjusted for mediator | 1.04 (0.97-1.12) | 1.01 (0.94-1.08) | 0.99 (0.93-1.07) | 1.01 (0.94-1.10) | 1.01 (0.94-1.09) |
| Mediator → dementia, adjusted for WHR | **0.90 (0.83-0.97)** | 0.98 (0.91-1.04) | 0.97 (0.89-1.04) | 1.03 (0.95-1.11) | 1.03 (0.96-1.10) |

Risk of dementia in relation to one standard deviation higher WHR or BMI and potential mediators measured in midlife or late-life. Hazard rate ratios (95% confidence intervals) were obtained from Cox proportional hazard models, with age as the underlying timescale. A hazard rate ratio above 1 indicates increased risk and below 1 indicates decreased risk of disease. It should be noted that only individuals with measures of both BMI or WHR and the potential mediator were included in the independent effect models above, leading to slight differences in adiposity estimates across models (number of individuals in each set of models are presented in the first row). All models were adjusted for sex and education. Models were stratified by study to allow for differences in the baseline hazard, and robust standard errors were applied to adjust for relatedness among twins. Models of lipid levels were additionally adjusted for fasting status. Bold numbers indicate significance at the $\alpha=0.05$ level.
BMI: body mass index; CRP: C-reactive protein; HDL: high-density lipoprotein; LDL: low-density lipoprotein; N: number of individuals; WHR: waist-hip ratio

Sex-stratified analyses showed a stronger effect of WHR in men (HRR 1.50, 95% CI 1.08-2.08) than women (HRR 1.13, 95% CI 0.88-1.45). Likewise, the effects of CRP, total cholesterol, and HDL-c, were stronger in men than women although estimates were generally not significant (Supplementary section, Table S8). Removing controls younger than 70 at end of follow-up had minimal effect on the estimates (Supplementary section, Table S9).

#### 3.2.2 Mediating effects of CRP and lipid levels on the association between adiposity and time to dementia

As the association between midlife WHR and dementia was stronger than that between midlife BMI and dementia, WHR was carried forward for mediation analyses. One SD higher WHR was associated with higher levels of CRP and triglycerides and lower levels of HDL-c, but not with total cholesterol or LDL-c (Table 3a; arrow a in Figure 1). In the AFT models, 1 SD higher WHR and total cholesterol were both associated with 2% shorter time to dementia onset, but the mediation decomposition indicated that the excess risk was driven by direct effects of WHR. It should be noted that total cholesterol does not meet the formal requirements of a mediator, as it was not associated with WHR. AFT models of WHR and the other biomarkers indicated no significant effects on time to dementia onset (Table 3a). This indicates that the association between midlife WHR and dementia risk goes mainly through direct effects of WHR rather than through CRP or lipid levels, i.e. through arrow c in Figure 1.

**Table 3:** Mediation models of the association between adiposity and dementia risk by CRP and lipid levels.

|  | CRP | Total cholesterol | HDL cholesterol | LDL cholesterol | Triglycerides |
| --- | --- | --- | --- | --- | --- |
| **a) <u>Midlife, WHR and potential mediators</u>** |  |  |  |  |  |
| **AFT model for the outcome** |  |  |  |  |  |
| WHR | 0.986 (0.971-1.003) | **0.981 (0.963-1.000)** | 0.989 (0.974-1.004) | 0.990 (0.975-1.005) | 0.986 (0.968-1.004) |
| Mediator | 1.008 (0.992-1.026) | **0.978 (0.961-0.996)** | 0.985 (0.969-1.001) | 0.996 (0.981-1.011) | 0.994 (0.975-1.013) |
| WHR*mediator | 1.000 (0.984-1.015) | 1.012 (0.997-1.026) | 0.993 (0.979-1.007) | 0.993 (0.978-1.008) | 1.002 (0.986-1.019) |
| **Linear model for the mediator** |  |  |  |  |  |
| WHR | **0.299 (0.273-0.324)** | -0.005 (-0.030-0.020) | **-0.296 (-0.318--0.274)** | 0.002 (-0.025-0.028) | **0.330 (0.306-0.355)** |
| **4-way decomposition of mediation** |  |  |  |  |  |
| Total excess relative risk | -0.011 (-0.029-0.006) | -0.017 (-0.035-0.001) | -0.002 (-0.020-0.015) | -0.007 (-0.025-0.010) | -0.015 (-0.033-0.003) |
| Controlled direct effect | -0.013 (-0.029-0.002) | **-0.019 (-0.038--0.001)** | -0.011 (-0.026-0.004) | -0.010 (-0.025-0.005) | -0.014 (-0.032-0.004) |
| Reference interaction | -0.000 (-0.004-0.004) | 0.002 (-0.001-0.005) | 0.002 (-0.002-0.006) | 0.003 (-0.004-0.010) | -0.001 (-0.004-0.005) |
| Mediated interaction | -0.000 (-0.005-0.004) | -0.000 (-0.000-0.000) | 0.002 (-0.002-0.006) | -0.000 (-0.000-0.000) | 0.001 (-0.005-0.006) |
| Pure indirect effect | 0.003 (-0.003-0.008) | 0.000 (-0.000-0.001) | 0.005 (-0.000-0.009) | -0.000 (-0.000-0.000) | -0.002 (-0.008-0.004) |
| **b) <u>Late-life, BMI and potential mediators</u>** |  |  |  |  |  |
| **AFT model for the outcome** |  |  |  |  |  |
| BMI | 1.004 (0.997-1.010) | **1.008 (1.001-1.014)** | **1.010 (1.004-1.017)** | **1.008 (1.001-1.014)** | **1.009 (1.002-1.016)** |
| Mediator | **1.008 (1.001-1.014)** | 1.003 (0.997-1.009) | **1.009 (1.002-1.016)** | 0.998 (0.992-1.004) | 0.994 (0.988-1.000) |
| BMI*mediator | 0.999 (0.993-1.006) | 1.001 (0.995-1.008) | **1.006 (1.000-1.012)** | 1.001 (0.994-1.008) | 0.996 (0.990-1.003) |
| **Linear model for the mediator** |  |  |  |  |  |
| BMI | **0.265 (0.241-0.289)** | **-0.063 (-0.085--0.041)** | **-0.272 (-0.293--0.251)** | **-0.058 (-0.084--0.033)** | **0.3031 (0.279-0.324)** |
| **4-way decomposition of mediation** |  |  |  |  |  |
| Total excess relative risk | 0.007 (-0.001-0.012) | 0.007 (-0.000-0.014) | 0.003 (-0.005-0.011) | **0.007 (0.000-0.014)** | 0.006 (-0.000-0.012) |
| Controlled direct effect | 0.003 (-0.003-0.010) | **0.008 (0.001-0.014)** | **0.010 (0.004-0.017)** | **0.008 (0.001-0.014)** | **0.009 (0.002-0.016)** |
| Reference interaction | -0.000 (-0.000-0.000) | -0.001 (-0.003-0.002) | **-0.003 (-0.006--0.000)** | -0.000 (-0.001-0.001) | 0.000 (-0.000-0.000) |
| Mediated interaction | -0.001 (-0.002-0.002) | -0.000 (-0.000-0.000) | **-0.002 (-0.003--0.000)** | -0.000 (-0.000-0.000) | -0.001 (-0.003-0.001) |
| Pure indirect effect | **0.001 (0.000-0.004)** | -0.000 (-0.001-0.000) | **-0.002 (-0.004--0.001)** | 0.000 (-0.000-0.000) | -0.002 (-0.004-0.000) |

Mediation models of one standard deviation higher WHR in midlife or BMI in late-life and dementia risk, by CRP, total cholesterol, HDL-cholesterol, LDL-cholesterol, and triglyceride levels. Time ratios in relation to one standard deviation higher BMI, WHR or potential mediator were obtained from accelerated failure time models with a Weibull distribution (the outcome model) with age as the timescale. A time ratio above 1 indicates longer disease-free time and thus a protective effect, while a time ratio below 1 indicates shorter disease-free time. Beta values for the association between BMI or WHR and the potential mediators were obtained from linear regression models (the mediator model). Analyses were adjusted for sex, education, and study. Models of lipid levels were additionally adjusted for fasting status. The total excess relative risk was decomposed into that explained by controlled direct effects of BMI or WHR, reference interaction, mediated interaction, and pure indirect effects of the mediator. Bold numbers indicate significance at the $\alpha=0.05$ level.
AFT: accelerated failure time; BMI: body mass index; CRP: C-reactive protein; HDL: high-density lipoprotein; LDL: low-density lipoprotein; WHR: waist-hip ratio

### 3.2 Late-life measures of adiposity and potential mediators in relation to dementia

#### 3.2.1 Independent and joint effects of adiposity, CRP, and lipid levels on the risk of dementia

In the full late-life sample, having 1 SD higher BMI was associated with lower dementia risk (HRR 0.92, 95% CI 0.86-0.99%), but no effect was seen for WHR (HRR 1.01, 95% CI 0.94-1.07). Including both BMI and WHR in the same model had little effect, resulting in HRR of 0.90 (95% CI 0.84-0.97) for BMI and 1.05 (95% CI 0.98-1.12) for WHR.

One SD higher CRP, but not lipid biomarkers, was associated with a lower risk of dementia (Table 2c-d). Jointly modelling BMI and CRP only slightly attenuated the estimates for both markers (Table 2c).

In sex-stratified analyses the inverse association between BMI and dementia was present mainly in women (HRR 0.90, 95% CI 0.83-0.98 in women, HRR 0.97, 95% CI 0.86-1.10 in men), but the effect of potential mediators did not markedly differ by sex (Supplementary section, Table S8).

#### 3.2.2 Mediating effects of CRP and lipid levels on the association between adiposity and time to dementia

BMI was selected for mediation analyses of late-life measures. One SD higher BMI was associated with higher levels of CRP and triglycerides, and lower levels of HDL-c, LDL-c, and total cholesterol (Table 3b). In the AFT models, 1 SD higher BMI, CRP, and HDL-c were significantly associated with a longer dementia free time (Table 3b).

The mediation decomposition of the BMI-CRP-dementia association indicated a significant pure indirect effect of CRP (Table 3b), which mediated 37% of the inverse association between BMI and dementia risk. This indicates that higher BMI is associated with higher CRP levels, which in part explains the inverse association between BMI and dementia, i.e. the effect goes through arrow a+b and c in Figure 1.

Decomposition of the BMI-HDL-c-dementia association indicated inconsistent mediation, with positive direct effects of BMI, together with negative interaction and mediation effects through HDL-c. As higher BMI is associated with lower HDL-c, which in turn is associated with shorter time to dementia, the inverse association between BMI and dementia is suppressed by BMI decreasing HDL-c levels. To better understand the relationship, the direct effect of BMI was tested with HDL-c levels fixed at -2 to 2 SD from the mean. Here, the direct effect of BMI was stronger with higher levels of HDL-c (Figure 2). Thus, higher late-life BMI was associated with lower dementia risk only if HDL-c remained high. No evidence of mediating effects from the other lipid biomarkers was present.

**Figure 2:**
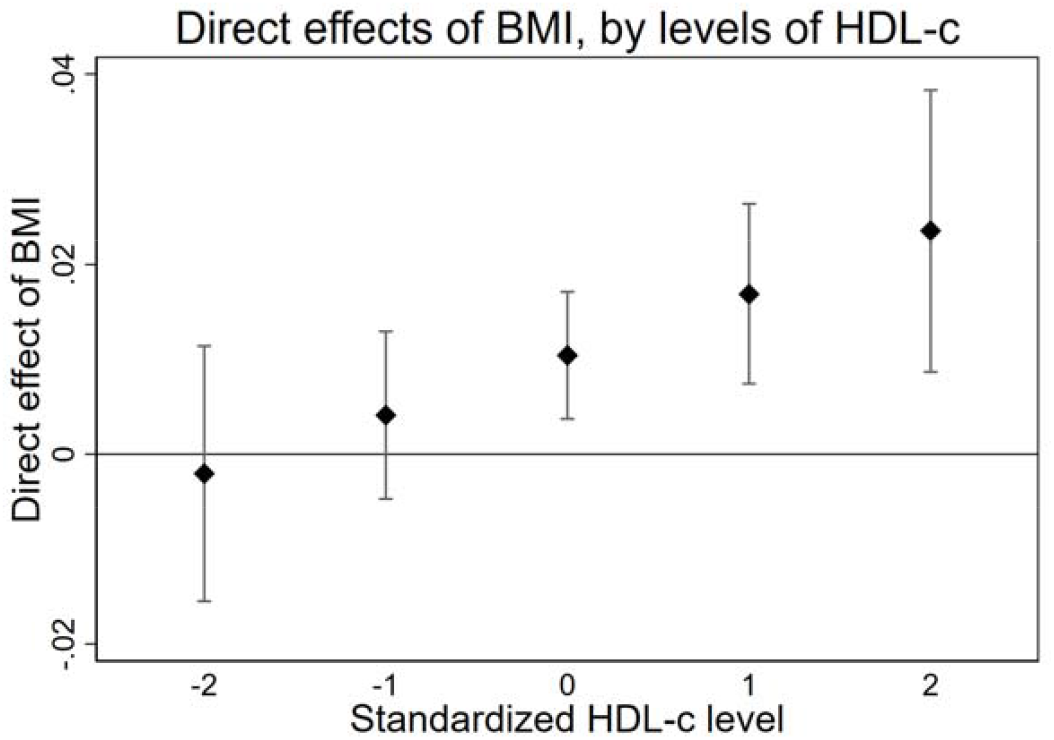
Excess relative risk of dementia due to direct effects of BMI, with HDL-c levels fixed at -2 to 2 standard deviations from the mean. Estimates were obtained from a mediation model with BMI as exposure, HDL-c as the mediator, and dementia as the outcome, with 4-way decomposition into direct effects, reference interaction, mediated interaction, and pure indirect effects. A linear regression model was used for the effect of BMI on HDL-c, and an accelerated failure time for the joint and interactive effects of BMI and HDL-c on time to dementia. The model was adjusted for age, sex, education, and study.

## 4. Discussion

Using publicly available summary statistics, we first used pathway analysis to guide selection of potential mediators, highlighting that genes influencing both adiposity and AD are involved in inflammation and lipid metabolism. Using individual level data, we then demonstrated that a higher WHR in midlife was strongly associated with dementia risk, and that the association was only slightly attenuated when adjusting for BMI. Mediation analyses indicated that the association was not explained by mediating effects of CRP, total cholesterol, HDL-c, LDL-c, or triglycerides. However, comparatively few events occurred in the midlife sample, and results should be interpreted with caution. In late-life, a higher BMI, but not WHR, was associated with lower risk of dementia, and the association was robust to adjustment for WHR. Mediation analyses indicated that part of the inverse association between late-life BMI and dementia risk was mediated by higher levels of CRP, where higher BMI is associated with higher CRP levels, which are in turn inversely associated with dementia. In addition, there was evidence of inconsistent mediation through HDL-c, where higher BMI was associated with lower HDL-c, which in turn was associated with higher risk of dementia, thus suppressing the inverse association between late-life BMI and dementia.

The age-specific effects of overweight on dementia risk are well established(1, 2), and the different effects of BMI and WHR seen in this study further highlight differences between midlife and late-life adiposity. In midlife, we found no evidence of mediating effects of inflammation or cholesterol levels, but rather, the effect appears to be driven mainly by other direct effects of WHR. Importantly, adjusting for BMI only slightly attenuated the effect of WHR. As mentioned, WHR reflects central adiposity, which indicates visceral fat storage and is more strongly linked to adverse health outcomes and metabolic dysfunction than gluteal adiposity(3). The association between midlife WHR and dementia was stronger among men, potentially because women generally have more gluteal fat compared to men, especially prior to menopause(24).

In late-life, BMI, but not WHR, was associated with dementia, indicating that the inverse association may indeed be driven by weight loss and not influenced by mechanisms related to body fat distribution. The reason for weight loss in preclinical dementia remains poorly understood, but evidence indicates it is a result of dementia pathology(25) rather than cognitive decline(26) or aging in general(27). Here, the association was present mainly among women, potentially mirroring more weight loss in women due to higher rates of disability and poor health compared to men, despite longer lifespans(28). Our results indicate that part of the inverse association is mediated through higher levels of CRP, a non-specific marker increased in both acute and chronic inflammation(29) linked to adiposity(30). The association between CRP and dementia also appears to have a paradoxical and age-specific pattern, similar to that of overweight and dementia. A meta-analysis of 8 prospective studies concluded that higher CRP levels are associated with increased risk of incident dementia(31), with stronger effects in studies with longer follow-ups. However, a meta-analysis of cross sectional studies shows that CRP levels are lower in individuals diagnosed with mild or moderate dementia, compared to controls(32), indicating that the association differs before and after disease onset. Thus, previous findings and our results indicate that the physiological processes leading to weight loss in preclinical dementia are linked to inflammation. Higher plasma HDL-c is inversely associated with dementia, both prospectively(33) and after disease onset(34), potentially through protection against cerebrovascular dysfunction(33). Our results demonstrated mediation and interaction effects through HDL-c, with a direct effect of higher BMI on dementia being present only when HDL-c levels remained at or above the mean. This may indicate that higher BMI is associated with lower risk of dementia only if metabolic function remains intact, highlighting the importance of considering adiposity together with metabolic health. Taken together, the late-life findings may mirror a lack of physiological homeostasis, seen as weight loss, loss of ability to mount an effective inflammatory response, and dysregulation of lipid metabolism, leading up to dementia.

### Strengths and limitations

This study first used summary statistics from large GWAS of AD, BMI, and WHR_adjBMI_ to identify biological pathways involved in both adiposity and dementia, thus highlighting potential mediating factors. The use of GWAS summary statistics, while powerful, comes with some limitations. Firstly, the GWAS of AD is substantially smaller than those for BMI and WHR_adjBMI_, and resulted in far fewer mapped genes. Secondly, the GWASs for BMI and WHR_adjBMI_ are based on study samples of a wide age-range and analyses are controlled for age. Hence, the results do not capture potential differences in genetic architecture of adiposity in midlife versus late-life. Taken together, this may have led to some relevant biological pathways being overlooked. The individual-level data are based on a well-characterized sample, with measures of both BMI, WHR, CRP, and lipid levels, together with dementia information both from the data collections and through linkage to nationwide disease registers. Most dementia diagnoses came from register information, and while the registers provides an opportunity to follow individuals far beyond the data collections, a substantial number of dementia diagnoses may be missed(16). In addition, while differential diagnosis is available from the register information, there is a large degree of misclassification(16). We therefore chose to study any dementia in the current studies, but cannot rule out differences in effects between dementia subtypes. It should be mentioned that a large part of the midlife sample is from the TwinGene study, which was conducted 2004-2008. As this results in only 8-12 years of follow-up, to a mean age of 69, it is important to highlight that many of the participants may have not yet developed or been diagnosed with dementia, and that those diagnosed may have comparatively early disease onset. Sensitivity analyses excluding controls aged <70 yielded very similar results as those from the main sample, but in light of this and the low number of dementia diagnoses, the midlife results should be interpreted with caution. A causal interpretation of mediation analysis assumes no unmeasured confounding between the exposure, mediator, and outcome. In addition, a mediator cannot be statistically distinguished from a confounder (reversed direction of arrow a in Figure 1) or collider (reversed direction of arrow b in Figure 1), but are only conceptually defined(35). Moreover, we used cross-sectional measures of adiposity and mediators, and the direction of effects between them cannot be examined. As adiposity, inflammation, and lipid metabolism are all complex biological systems, with long-term effects and unclear links to dementia, we make no causal claims based on these results.

## Conclusions

In conclusion, we demonstrated that a higher WHR in midlife is linked to increased risk of dementia, and that this association is not explained by CRP or blood lipid levels. In late-life, a higher BMI is associated with lower risk of dementia, with a substantial proportion of the association mediated through CRP levels while the association is suppressed by lower levels of HDL-c. Taken together, this strengthens the difference between adiposity in midlife and late-life, and shows that central adiposity in midlife may be an important target for disease prevention, while weight loss in late-life may be a warning sign of ill health and warrant observation of additional signs of dementia pathology.

## Data Availability

The data are held by the Swedish Twin Registry, and can be applied for through the link below.
Codes, logs, and outputs are available on: https://github.com/ik-karlsson/Adiposity_dementia_mediation

https://ki.se/en/research/swedish-twin-registry-for-researchers

## Acknowledgements

This work was supported by the Strategic Research Program in Epidemiology at Karolinska Institutet; the Swedish Research Council for Health, Working Life and Welfare (2018-01201); the Swedish Research Council (2016-03081); and the National Institutes of Health (R01 AG060470).

We acknowledge the Swedish Twin Registry for access to data. The Swedish Twin Registry is managed by Karolinska Institutet and receives funding through the Swedish Research Council under the grant no. 2017-00641.

The STR sub-studies were supported by the National Institutes of Health (grants R01 AG10175, R01 AG08724, R01 AG08861, R01 AG028555, and U01 DK066134), the MacArthur Foundation Research Network on Successful Aging, the Axel and Margaret Ax:son Johnsons Foundation, the Swedish Research Council, the Swedish Foundation for Health Care Sciences and Allergy Research, and the Swedish Council for Working Life and Social Research (2013-2292).

## Potential conflicts of interest

None of the authors have any conflicts of interest to report.

## Notes

### Competing Interest Statement

The authors have declared no competing interest.

### Author Declarations

Regional Ethical Review Board in Stockholm

